# Recurrence of SARS-CoV-2 PCR positivity in COVID-19 patients: a single center experience and potential implications

**DOI:** 10.1101/2020.05.06.20089573

**Authors:** Jia Huang, Le Zheng, Zhen Li, Shiying Hao, Fangfan Ye, Jun Chen, Xiaoming Yao, Jiayu Liao, Song Wang, Manfei Zeng, Liping Qiu, Fanlan Cen, Yajing Huang, Tengfei Zhu, Zehui Xu, Manhua Ye, Yang Yang, Guowei Wang, Jinxiu Li, Lifei Wang, Jiuxin Qu, Jing Yuan, Wei Zheng, Zheng Zhang, Chunyang Li, John C. Whitin, Lu Tian, Henry Chubb, Kuo-Yuan Hwa, Hayley A. Gans, Scott R. Ceresnak, Wei Zhang, Ying Lu, Yvonne A. Maldonado, Qing He, Zhaoqin Wang, Yingxia Liu, Doff B. McElhinney, Karl G. Sylvester, Harvey J. Cohen, Lei Liu, Xuefeng B. Ling

**Affiliations:** National Clinical Research Center for Infectious Disease, The Second Affiliated Hospital of Southern University of Science and Technology, Shenzhen, Guangdong Province, China; Department of Surgery, Stanford University School of Medicine, Stanford, CA, United States; Department of Cardiothoracic Surgery, Stanford University School of Medicine, Stanford, CA, United States; Clinical and Translational Research Program, Betty Irene Moore Children’s Heart Center, Lucile Packard Children’s Hospital, Palo Alto, CA, United States; West China Hospital, Sichuan University, Chengdu, China; Department of Bioengineering, University of California at Riverside, Riverside, CA, USA; Biomedical Big Data Center, West China Hospital, Sichuan University, Chengdu, China; Medical Big Data Center, Sichuan University, Chengdu, China; Department of Pediatrics, Stanford University School of Medicine, Stanford, CA, United States; Department of Biomedical Data Science, Stanford University, Stanford, CA, United States; Department of Medicine, The University of Hong Kong, Hong Kong SAR, China; Department of Health Research and Policy, Stanford University School of Medicine, Stanford, CA, United States

## Abstract

**IMPORTANCE:** How to appropriately care for patients who become PCR-negative for severe acute respiratory syndrome coronavirus 2 (SARS-CoV-2) is still not known. Patients who have recovered from coronavirus disease 2019 (COVID-19) could profoundly impact the health care system if a subset were to be PCR-positive again with reactivated SARS-CoV-2.

**OBJECTIVE:** To characterize a single center COVID-19 cohort with and without recurrence of PCR positivity, and develop an algorithm to identify patients at high risk of retest positivity after discharge to inform health care policy and case management decision-making.

**DESIGN, SETTING, AND PARTICIPANTS:** A cohort of 414 patients with confirmed SARS-CoV-2 infection, at The Second Affiliated Hospital of Southern University of Science and Technology in Shenzhen, China from January 11 to April 23, 2020.

**EXPOSURES:** Polymerase chain reaction (PCR) and IgM-IgG antibody confirmed SARS-CoV-2 infection.

**MAIN OUTCOMES AND MEASURES:** Univariable and multivariable statistical analysis of the clinical, laboratory, radiologic image, medical treatment, and clinical course of admission/quarantine/readmission data to develop an algorithm to predict patients at risk of recurrence of PCR positivity.

**RESULTS:** 16.7% (95CI: 13.0%-20.3%) patients retest PCR positive 1 to 3 times after discharge, despite being in strict quarantine. The driving factors in the recurrence prediction model included: age, BMI; lowest levels of the blood laboratory tests during hospitalization for cholinesterase, fibrinogen, albumin, prealbumin, calcium, eGFR, creatinine; highest levels of the blood laboratory tests during hospitalization for total bilirubin, lactate dehydrogenase, alkaline phosphatase; the first test results during hospitalization for partial pressure of oxygen, white blood cell and lymphocyte counts, blood procalcitonin; and the first test episodic Ct value and the lowest Ct value of the nasopharyngeal swab RT PCR results. Area under the ROC curve is 0.786.

**CONCLUSIONS AND RELEVANCE:** This case series provides clinical characteristics of COVID-19 patients with recurrent PCR positivity, despite strict quarantine, at a 16.7% rate. Use of a recurrence prediction algorithm may identify patients at high risk of PCR retest positivity of SARS-CoV-2 and help modify COVID-19 case management and health policy approaches.

**Key Points:** *Question:* What are the characteristics, clinical presentations, and outcomes of COVID-19 patients with PCR retest positivity after resolution of the initial infection and consecutive negative tests? Can we identify recovered patients, prior to discharge, at risk of the recurrence of SARS-CoV-2 PCR positivity?

*Findings:* In this series of 414 COVID-19 inpatients discharged to a designated quarantine center, 69 retest positive (13 with 2 readmissions, and 3 with 3 readmissions). A multivariable model was developed to predict the risk of the recurrence of SARS-CoV-2 PCR positivity.

*Meaning:* Rate and timing of the recurrence of PCR positivity following strict quarantine were characterized. Our prediction algorithm may have implications for COVID-19 clinical treatment, patient management, and health policy.

## Introduction

Given the sudden emergence and rapid community transmission of severe acute respiratory syndrome coronavirus 2 (SARS-CoV-2) being observed worldwide, a strategy of social distancing and shelter in place has been widely adopted in an effort to curb the spread of COVID-19 across space and time^1,2^. The quarantining of patients testing positive for SARS-CoV-2 virus is considered mandatory in order to prevent continued viral spread (contagion). In last 6 months, many of COVID-19 patients have since clinically recovered and been discharged from the hospital, but it remains unclear the degree to which patients with COVID-19 (clinical symptoms and PCR test positivity) remain contagious and or at risk for disease relapse. The rising concern is that COVID-19 discharged patients may be at risk of viral reactivation to infect others as asymptomatic carriers, or be re-infected themselves. In an attempt to better understand these concerns, varying quarantine strategies have been implemented during the transition of COVID-19 recovering patients from healthcare to non-healthcare settings in this current pandemic.

Recently, the early experiences of 116 cases confirmed by nasopharyngeal swab testing, potentially resulting from either “reactivated” or “re-infected” SARS-CoV-2, was reported in South Korea^3,4^. In response, the World Health Organization (WHO) commented that there is currently “no evidence” demonstrating that people who have recovered from the coronavirus are not at risk of re-infection^5^.

However, limited information is available regarding viral shedding kinetics and live virus isolation. Variability in PCR methodology will result in different thresholds of the assay for RNA detection, but in one study the SARS-CoV-2 RNA threshold upon PCR testing needs to be greater than 10^6^ copies per sample^6^. To date, there have been no reports of live virus isolation from patients of PCR retest positivity. Importantly, though, pathological evidence of likely viable SARS-CoV-2 virus within pneumocytes was recently obtained from an individual who died unexpectedly from cardiac arrest after showing clinical recovery and three consecutive negative PCR nasopharyngeal swabs.^7^

In order to assist with pandemic management, a better understanding of the recurrence of SARS-CoV-2 and associated potential infectivity in the setting of strict quarantine is critical. Similarly, to assist in managing individuals, setting quarantine strategies and adjudicating limited healthcare resources, prediction models are needed to better define the risk, timing, and relevance of viral PCR retest positivity. Unanswered questions include: time between nasopharyngeal swab test negative and length of effective quarantine, and how infectious is an infected person who recurred with PCR positivity after testing negative. Pragmatic models should seek to define when recovered patients can be infectious, including in the cohort of individuals who recur with PCR positivity relative to testing interval. Thus, limited resources can be concentrated on the isolation of these potential SARS-CoV-2 carriers, with immediate benefits for the patient, the population and the healthcare system.

This study characterizes a single center cohort of consecutive patients with COVID-19 who were followed after recovery and PCR negativity and shown to have one or more recurrent positive PCR result despite continued quarantine. The primary objective was to describe the kinetics of SARS-CoV-2 PCR in a large cohort of infected individuals and better understand the relevance of recurrent positive results. The secondary objective was to develop a prediction algorithm to identify patients at high risk of the recurrent PCR positivity and provide practical data that may impact medical operation, health care policy, and case management.

## Methods

### Study design and participants

The study cohort included consecutive COVID-19 patients admitted to The Second Affiliated Hospital of Southern University of Science and Technology, Shenzhen, China since January 11, 2020. The last follow up date was April 23, 2020. All discharged COVID-19 patients were subjected to strict quarantine at a designated center for 14 days. SARS-CoV-2 quantitative reverse transcription polymerase chain reaction (qRT-PCR) RNA testing was performed every 3 to 5 days during both hospitalization and quarantine. Follow-up at home quarantine was mandated for an additional 14 days with weekly SARS-CoV-2 qRT-PCR testing. Upon positive nasopharyngeal swab testing, according to the local health policy, these patients were immediately readmitted back to the hospital (Supplementary Figure 1). This study was approved by the Ethics Committee of the Second Affiliated Hospital of Southern University of Science and Technology. Written informed consent was obtained from all patients.

Demographic features, comorbidities, clinical symptoms, vital signs, laboratory findings and treatments during the first hospitalization were collected. Sequentially from admission, nasopharyngeal swab testing was performed every 3 days during hospitalization. Reported treatment information includes medicines, intensive care unit (ICU) admissions, and respiratory support and ventilation usage.

### Admission and discharge criteria of COVID-19 pneumonia

Diagnosis, disease severity, treatment and follow-up criteria for COVID-19 infection were based on the preliminary diagnosis and treatment protocols (6th edition) from the National Health Commission of China^8^. Diagnostic criteria for COVID-19 pneumonia included epidemiological (demographics and comorbidities) history, typical clinical manifestations (fever, respiratory symptoms) and laboratory diagnosis. Pulmonary respiratory syndrome severity was classified into 4 categories: (1) mild: mild respiratory symptoms, no imaging findings of pneumonia; (2) moderate: fever, respiratory symptoms, imaging findings of pneumonia; (3) severe: shortness of breath, respiratory rate >30 breaths/min, systemic oxygen saturation <93% at rest on room air, ratio of the systemic arterial partial pressure of oxygen to the fraction of oxygen in inspired air ≤300mmHg, or >50% progression of radiologic pulmonary lesions over 24 to 48 hours; (4) critical: needing mechanical ventilation, extracorporeal membrane oxygenation, or other organ support therapy in the ICU. The discharge criteria included: being afebrile for at least three days, resolved respiratory symptoms, improvement of radiological abnormities in CT or X-ray, and two consecutive negative SARS-Cov-2 qRT-PCR tests sampled >1 day apart. All patients were discharged under strict monitoring conditions: patients were kept for 14 days at a designated center followed by another 14 days at home, in quarantine, and all discharged patients were tested with repeated routine SARS-CoV-2 qRT-PCR detections in nasopharyngeal swab samples. According to the local quarantine policy, patients with a positive SARS-CoV-2 qRT-PCR nasopharyngeal test were immediately readmitted back to the hospital.

### SARS-CoV-2 tests

Early morning nasopharyngeal swabs (from January 12, 2020) and anal swabs (from February 2, 2020) were analyzed every 3 days during the hospitalization, every 3 to 5 days during mandated quarantine at a designated center, and weekly during quarantine at home. Bronchoalveolar lavage washing was sampled from patients with severe illness or undergoing mechanical ventilation. Total RNA was extracted from the clinical specimens using the QIAamp RNA Viral Kit (Qiagen, Heiden, Germany). A qRT-PCR Test Kit (product code. GZ-D2RM25, Shanghai GeneoDx Biotech Co., Ltd) targeting the ORF1ab and N genes of SARS-CoV-2 was used. A cycle threshold (Ct) value less than 37 was interpreted as positive for SARS-CoV-2 RNA^9^.

SARS-CoV-2 antibody Chemiluminescent microparticle immunoassay (CMIA) kit (Innodx, Xiamen, China; catalog no. Gxzz 20203400198) was used to detect SARS-CoV-2 IgM and IgG in plasma.

### Outcome

Patients who had a positive nasopharyngeal swab test during post-discharge follow-up and were readmitted to hospital were defined as ‘case’. Patients who did not have positive results of nasopharyngeal swab test after discharge were analyzed as ‘control’ patients.

### Statistical analysis and modelling to predict recurrence of PCR positivity

Features including demographics, comorbidities, symptoms, vital signs, laboratory findings, and treatments were assembled for modelling. Univariable analysis was performed on z-score-normalized features, and logistic regression was used to calculate the odds ratios and *P* values for feature filtering. For multivariate model building, a gradient boosting tree algorithm XGBoost was used for constructing a multivariable prediction model^10–16^. The baseline learner is the classification and regression tree and the number of trees is selected via cross-validation to avoid over-fitting. The derived model score ranged from 0 to 100 describing the probability of recurrence after discharge. The recurrence prediction model was evaluated using area under the receiver-operating-characteristic curve (ROC AUC), sensitivity, and specificity from the 10-fold cross-validations. Statistical analyses were performed using R software (version 3.5.1).

## Results

### Baseline characteristics

The study comprised a total of 417 consecutive patients admitted to the hospital with COVID-19 who were categorized to have mild (N=16), moderate (N=309), severe (N=73), or critical (N=19) conditions of pulmonary respiratory syndrome. Death occurred 3 of 417 (0.7%) patients during initial hospitalization. Of the remaining 414 patients alive, 69 [16.7% (95CI: 13.0%-20.3%), case] patients were with recurrence of nasopharyngeal swab PCR positivity and had ≥1 readmission(s). Demographics, clinical data, PCR data, and outcomes from the first hospitalization are summarized (Table 1, Table 2 and Supplementary Figure 2). Statistically significant differences between case and control patients were observed for patient age, body mass index, and clinical severity of pulmonary respiratory syndrome during the first hospitalization. Case patients were generally younger than the controls *(P* value <0.001). The majority (93%) of case patients had mild or moderate pulmonary respiratory syndrome at the first admission, and had respiratory symptoms including cough and increased sputum at the readmission of PCR positivity. Only 2 of the 69 patients, negative for other non-COVID-19 infections, were febrile with typical clinical manifestations satisfying the first admission criteria.

**Table 1.** Demographics and baseline characteristics of COVID-19 patients with (i.e. the case group) and without (i.e. the control group) recurrence of SARS-CoV-2 PCR positivity.

| Characteristics | Control, N=345 | Case, N=69 | P value |
| --- | --- | --- | --- |
| **Age, n (%)** |  |  | <0.001 |
| 0-29 yrs | 47 (13.6) | 23 (33) |  |
| 30-54 yrs | 164 (47.5) | 34 (49) |  |
| 55-86 yrs | 134 (38.8) | 12 (17) |  |
| **Male, n (%)** | 167 (48.4) | 28 (41) | 0.2 |
| **BMI, kg/m<sup>2</sup>** | 23.2 (21.3, 25.6) | 21.9 (20.0, 24.5) | 0.03 |
| **COVID-19 severity at 1<sup>st</sup> admission, n (%)** |  |  | 0.008 |
| Mild | 13 (3.8) | 3 (4) |  |
| Moderate | 248 (71.9) | 61 (88) |  |
| Severe | 68 (19.7) | 5 (7) |  |
| Critical | 16 (4.6) | 0 (0) |  |
| **Clinical history, n (%)** |  |  |  |
| Hypertension | 71 (20.6) | 14 (20) | 0.9 |
| Diabetes | 31 (9.0) | 3 (4) | 0.2 |
| Coronary heart disease | 25 (7.2) | 1 (1) | 0.1 |
| Cancer | 6 (1.7) | 0 (0) | 0.6 |
| Chronic lung disease | 14 (4.1) | 2 (3) | 0.9 |
| Chronic liver disease | 11 (3.2) | 2 (3) | 0.9 |

**Table 2.** Clinical characteristics, laboratory tests, treatments, and outcomes of COVID-19 patients with (i.e. the case group) and without (i.e. the control group) recurrence of SARS-CoV-2 PCR positivity during the 1^st^ hospitalization. Data are presented in form of n (%) or median (IQR), unless otherwise stated. For statistical analyses, the Mann-Whitney U test was used to compare continuous variables and Fisher’s exact test was performed to compare categorical variables between groups.

| Characteristics | Control, N=345 | Case, N=69 | P Value |
| --- | --- | --- | --- |
| **Symptoms, n (%)** |  |  |  |
| Fever | 228 (66.1) | 43 (62) | 0.6 |
| Cough | 33 (9.6) | 28 (41) | <0.0001 |
| Sputum | 15 (4.3) | 15 (22) | <0.0001 |
| Dizziness | 9 (2.6) | 3 (4) | 0.4 |
| Headache | 6 (1.7) | 7 (10) | 0.002 |
| Nasopharyngeal soreness | 14 (4.1) | 9 (13) | 0.007 |
| Shortness of breath | 2 (0.6) | 1 (1) | 0.9 |
| Tightness | 9 (2.6) | 4 (6) | 0.3 |
| Bloating | 2 (0.6) | 0 (0) | 0.9 |
| Diarrhea | 8 (2.3) | 3 (4) | 0.4 |
| Fatigue | 26 (7.5) | 8 (12) | 0.2 |
| Chest pain | 2 (0.6) | 1 (1) | 0.4 |
| Muscle or body aches | 49 (14.2) | 2 (3) | 0.008 |
| Chills | 5 (1.4) | 5 (7) | 0.02 |
| Nausea and vomiting | 3 (0.9) | 1 (1) | 0.5 |
| **Imaging feature, n (%)** |  |  |  |
| Lung consolidation | 68 (19.7) | 11 (16) | 0.5 |
| Ground-glass opacity | 282 (81.7) | 60 (87) | 0.4 |
| Pulmonary infiltration | 255 (73.9) | 58 (84) | 0.2 |
| Pleural effusion | 13 (3.8) | 2 (3) | 0.7 |
| **Medication Treatment, n (%)** |  |  |  |
| Methylprednisolone | 91 (26.4) | 8 (12) | 0.006 |
| Immunoglobulin | 97 (28.1) | 8 (12) | 0.002 |
| Tocilizumab | 9 (2.6) | 0 (0) | 0.2 |
| Oseltamivir | 54 (15.7) | 7 (10) | 0.3 |
| Ribavirin | 71 (20.6) | 9 (13) | 0.1 |
| Interferon | 287 (83.2) | 54 (78) | 0.3 |
| Lopinavir/Ritonavir | 271 (78.6) | 49 (71) | 0.2 |
| Arbidol | 102 (29.6) | 11 (16) | 0.03 |
| Favipiravir | 32 (9.3) | 3 (4) | 0.2 |
| Hydroxychloroquine sulfate | 21 (6.1) | 5 (7) | 0.9 |
| Antibiotics | 69 (20) | 7 (10) | 0.04 |

*Table 2 continues in next column* Continued from previous column
| Characteristics | Control, N=345 | Case, N=69 | P Value |
| --- | --- | --- | --- |
| **Supporting treatment, n (%)** |  |  |  |
| High-flow nasal cannula oxygen therapy | 16 (4.6) | 1 (1) | 0.3 |
| Non-invasive ventilation | 27 (7.8) | 0 (0) | 0.02 |
| Invasive ventilation | 16 (4.6) | 0 (0) | 0.05 |
| Extracorporeal membrane oxygenation | 2 (0.6) | 0 (0) | 0.9 |
| Continuous renal replacement treatment | 5 (1.4) | 0 (0) | 0.9 |
| **Blood routine, n (%)** |  |  |  |
| Hemoglobin, <13.7 g/dL (male)<br><11.9 g/dL (female) | 31 (9.0) | 4 (6) | 0.5 |
| Total white blood cell count |  |  |  |
| <3.5 × 10<sup>9</sup>/L | 58 (16.8) | 8 (12) | 0.4 |
| >9.5 × 10<sup>9</sup>/L | 11 (3.2) | 2 (3) | 0.9 |
| Lymphocyte count, <1.1 × 10<sup>9</sup>/L | 103 (29.9) | 12 (17) | 0.04 |
| Neutrophil count, >6.3 × 10<sup>9</sup>/L | 16 (4.6) | 2 (3) | 0.7 |
| Platelet count, <125 × 10<sup>9</sup>/L | 29 (8.4) | 8 (12) | 0.4 |
| **Blood biochemistry, n (%)** |  |  |  |
| Sodium, <135 mmol/L | 21 (6.1) | 4 (6) | 0.9 |
| Potassium, <3.5 mmol/L | 29 (8.4) | 4 (6) | 0.6 |
| Urea, >9.5 mmol/L | 3 (0.9) | 1 (1) | 0.5 |
| Creatinine, >111, μmol/L | 6 (1.7) | 0 (0) | 0.6 |
| Albumin, <40 g/L | 51 (14.8) | 5 (7) | 0.1 |
| ALT, >45 U/L | 27 (7.8) | 6 (9) | 0.8 |
| AST, >45 U/L | 36 (10.4) | 1 (1) | 0.02 |
| Lactate dehydrogenase, >250 U/L | 113 (32.8) | 12 (17) | 0.01 |
| Creatine kinase, >310 U/L | 7 (2.0) | 2 (3) | 0.6 |
| **Infection-related biomarkers, n (%)** |  |  |  |
| Erythrocyte sedimentation rate, >20 mm/h | 151 (43.8) | 22 (32) | 0.08 |
| Interleukin 6, >7 p/mL | 97 (28.1) | 11 (16) | 0.04 |
| Procalcitonin |  |  | 0.04 |
| 0.1 ng/mL | 335 (97.1) | 61 (88) |  |
| ≥0.1 to <0.25 ng/mL | 10 (2.9) | 4 (6) |  |
| ≥0.25 to ≤0.5 ng/mL | 0 (0) | 1 (1) |  |
| >0.5 ng/mL | 0 (0) | 1 (1) |  |
| C-reactive protein, >8 mg/L | 151 (43.8) | 21 (30) | 0.04 |

Table 2 continues in next column *Continued from previous column*
| **Characteristics** | **Control, N=345** | **Case, N=69** | **P Value** |
| --- | --- | --- | --- |
| **Coagulation function, n (%)** |  |  |  |
| Prothrombin time, ≥16 s | 3 (0.9) | 1 (1) | 0.5 |
| D-dimer |  |  | 0.5 |
| ≤0.5µg/mL | 265 (77.0) | 55 (80) |  |
| >0.5 and ≤1µg/mL | 58 (16.8) | 8 (12) |  |
| >1µg/mL | 22 (6.4) | 4 (6) |  |
| **SARS-CoV-2 antibody at discharge <sup>a</sup>** |  |  | 0.23 |
| Having IgG/IgM tested, n | 114 (33.0) | 40 (58) |  |
| IgG positive, n (%) | 113 (99.1) | 40 (100) |  |
| IgM positive, n (%) | 55 (48.2) | 30 (75) |  |
| **Clinical Outcomes** |  |  |  |
| ICU admission | 34 (9.9) | 0 (0) | 0.002 |
| Length of hospitalization | 20 (12) | 20 (12) | 0.5 |

<sup>a</sup> IgG/IgM tests were performed at discharge since February 12, 2020.

### Patient clinical history: admission, discharge, quarantine, and readmission

The timeline of clinical events including admission, discharge, quarantine and readmission are summarized (Figure 1, cases are divided into subgroups according to number of readmissions; Supplementary Figure 3). A total of 69 patients were re-test positive for SARS-CoV-2 RNA during the follow up period. The median time (days) from new onset of symptoms to either the first positive nasopharyngeal swab PCR test after admission or PCR test negative after treatment was 3 or 12 days, respectively. Distribution of time (days) from new onset of symptoms to either the first positive swab test or the first discharge was analyzed (Supplementary Figure 3 and Supplementary Figure 4).

**Figure 1.**
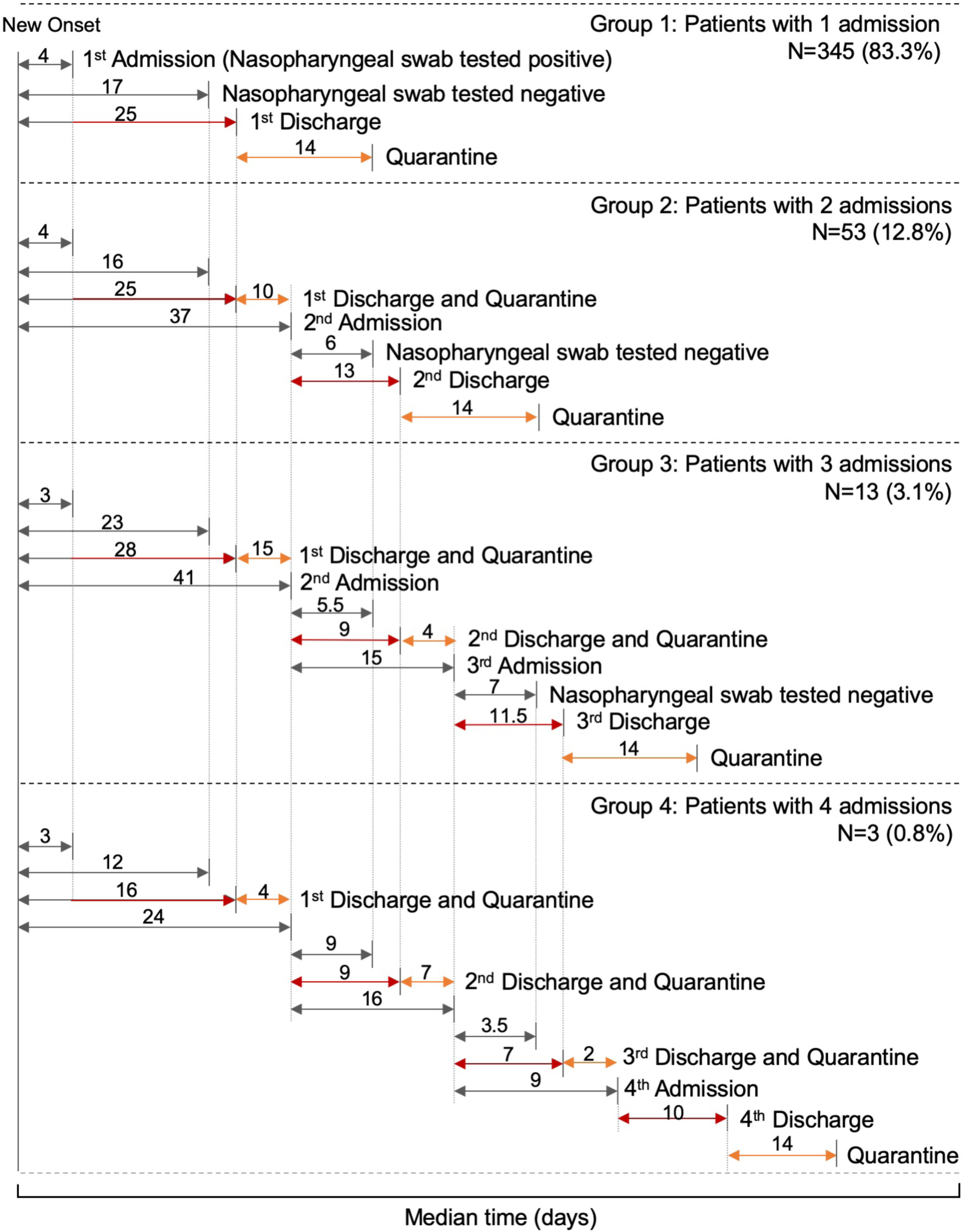
This timeline summarizes the median duration (days) from the onset of symptoms to clinical events and recurrence of nasopharyngeal swab PCR positivity. Patients are grouped by the number of (re)admissions. Clinical events include admission, nasopharyngeal swab tested negative, discharge, and quarantine ended due to either retest positive or release to home.

The 69 case patients had 1 (N=53), 2 (N=13), or 3 (N=3) readmissions for positive nasopharyngeal swab re-tests, most of whom had mild/moderate disease during the first admission (Figure 1). As of April 23^rd^, 67 of the 69 total cases have been discharged following two consecutive negative SARS-CoV-2 swab tests, while 2 patients remained within their second hospitalization. During the second period of post-discharge observation, 16 patients tested positive once again for SARS-CoV-2 RNA, indicating a median time of 8.5 days from test negative to retest positive, demonstrating a shorter inter-episode period than the first recurrence. Three patients tested positive for the fourth time following three quarantine periods with a median time of only 5.5 days from prior test negative to retest positive.

Recurrence of PCR positivity was analyzed as a function of time (days) between the first nasopharyngeal swab test negative during the first hospitalization and retest positive during the follow up period (Figure 2A, median 19 days, range 6-52 days). Within the case group, 70% retested positive within 5-25 days after the first negative test, with a peak occurring at 10-15 days (22%).

**Figure 2.**
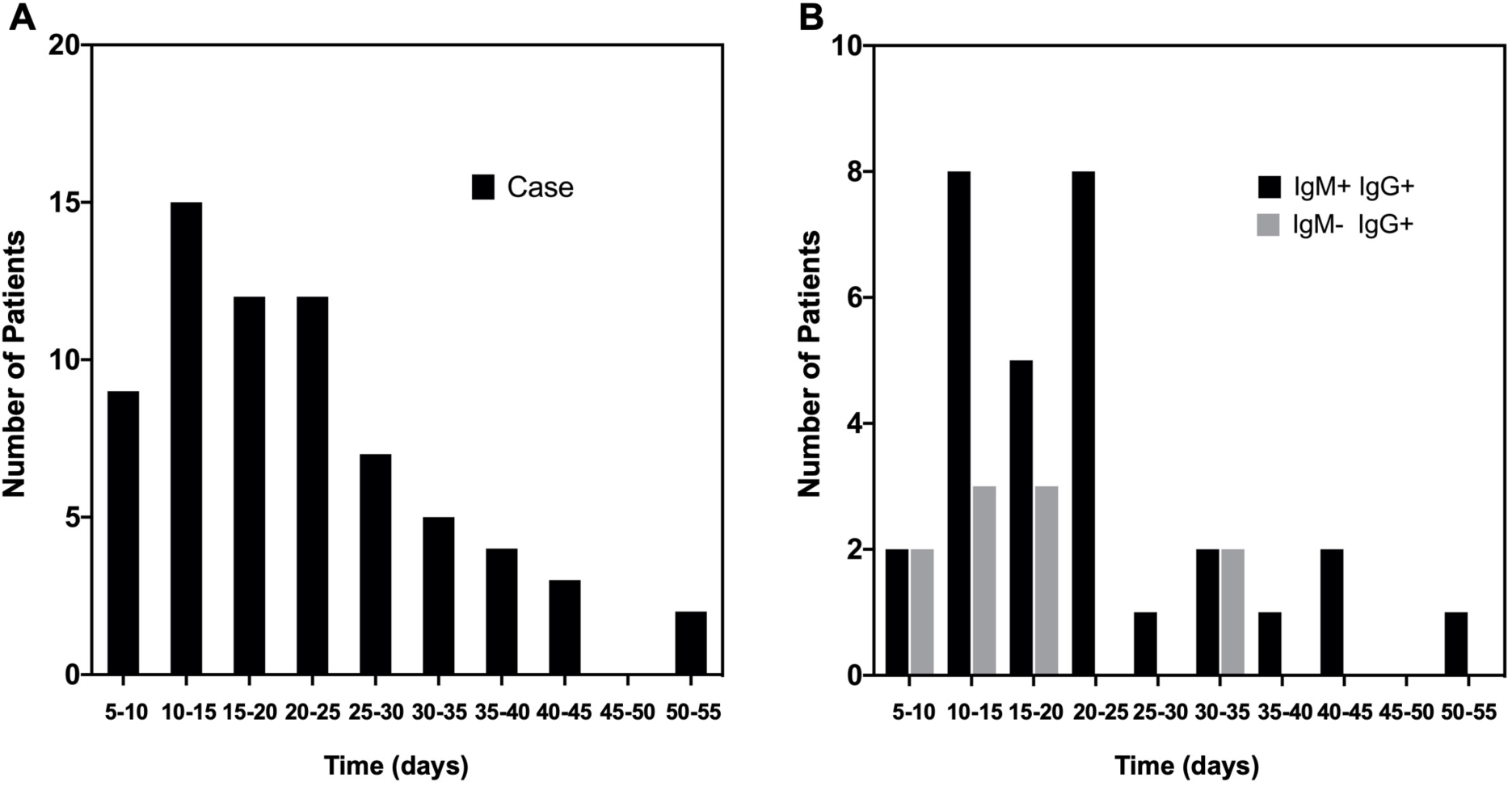
These column charts show the distribution of patients according to the duration (days) from the first negative consecutive nasopharyngeal swab test to the first positive retest during strict post-discharge quarantine. A) COVID-19 patients with recurrence of PCR positivity (the case group). B) The subset of case group (N=40) who had IgM and IgG antibody testing performed at the initial discharge. All case patients with the antibody tests were IgG positive.

### Serology results in case and control patients

Of note, a subset of 154 patients had IgG/IgM antibody testing at the initial discharge, among which 85 and 153 were IgG and IgM positive, respectively. 1/154 had repeated negative antibody tests (N=5) of both IgM and IgG against SARS-CoV-2. This suggests that convalescent patients, including aging populations^6^, may fail to develop SARS-CoV-2 specific IgM and IgG. Of the 154 patients tested, 40 (100%) of the case group were IgG positive, and 30 (75%) were IgM positive (Figure 2B and Supplementary Figure 5).

### Predicting recurrence of PCR positivity at discharge: model development and performance

The model was built with 69 cases and 345 controls. Eighteen clinical factors were selected based on *P* values and utilized by the XGBoost algorithm for the final model (Supplementary Table 1A and Supplementary Text 1).

The prediction model displayed an overall AUC of 0.786 based on 10-fold cross-validation (Supplementary Table 1B). To determine the accuracy and demonstrate the utility of the model to predict a future recurrence of PCR positivity, an analysis of ‘days to PCR positivity’ of high-risk patients was under-taken. This analysis supports our hypothesis that our prediction is feasible and may give actionable information at the time of the first of the consecutive nasopharyngeal swab negative tests during the hospitalization (Figure 3) or at the discharge time (Supplementary Figure 6).

**Figure 3.**
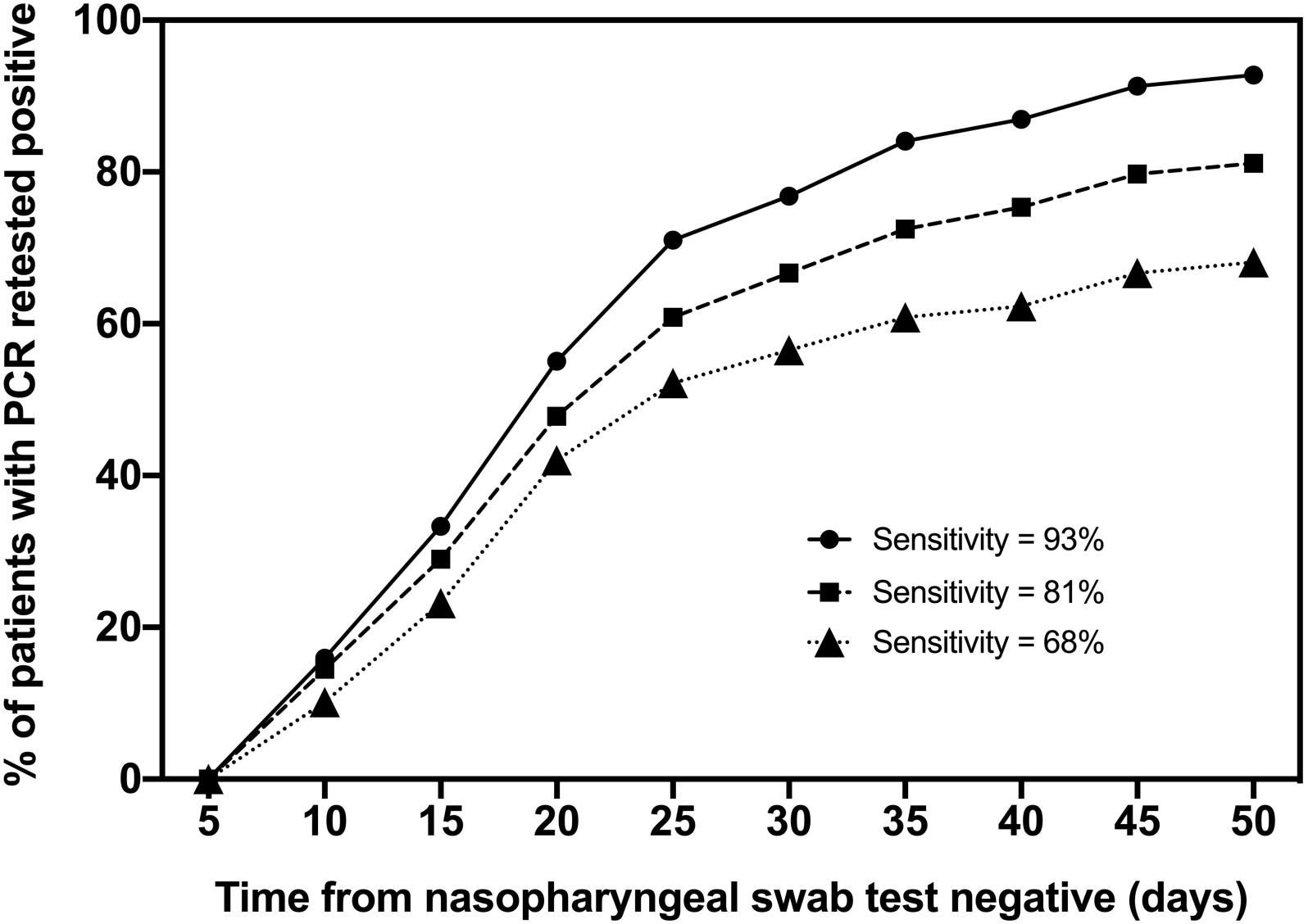
High-risk patients had nasopharyngeal swab positive retests and were readmitted to hospital. The X-axis indicates the duration of time (days) from the first negative nasopharyngeal swab test to the first positive retest during strict post-discharge quarantine. The Y-axis indicates the percentage of high-risk patients who were retested positive and readmitted to hospital within the specified duration of time after the first negative test. Three thresholds for high-risk patients were applied, giving an overall sensitivity of 93%, 81%, and 68%, respectively.

## Discussion

Given the rapidly evolving COVID-19 pandemic, it is critical to develop an understanding of the recurrence of PCR positivity relative to severity of clinical presentation and viral particle detection. The current study is important since the data show that a significant percentage of infected and recovered individuals will have recurrent PCR NP swab positivity despite strict quarantine. To our knowledge, this study is among the first to conduct a comprehensive assessment of PCR kenetics with regular serial viral RNA testing in the setting of strict quarantine. We also propose a case finding model to identify patients at high risk for COVID-19 viral retest positivity to assist in the determination of healthcare system utilization and health policy planning.

The 16.7% nasopharyngeal swab retest positive rate has not been previously reported. Given the mandated quarantine for discharged patients at a designated center for a minimum of 14 days followed by another 14 days at home (total 28 days of strict social distancing, shelter-in-place), the observed overall rate and multiple recurrence of positive viral testing are unlikely to be from re-infection. This contrasts with the recently case reports in South Korea^4,5^, wherein recovered patients were home quarantined and recurrence of PCR positivity there could have arisen from either dormant virus reactivation or re-infection.

COVID-19 patients with multiple recurrence of PCR positivity have not been previously reported. Thirteen and three cases recurred two and three times with PCR positivity, respectively, making it likely that the re-emergence of PCR positivity is due to cycling between dormancy and reactivation of SARS-CoV-2, and/or the resurfacing of the virus from the lower tract to the upper tract of the respiratory system. However, it must be noted that the positive SARS-CoV-2 test does not equate with infectivity. Since the current standard for SARS-CoV-2 test positivity is predicated on viral load detection by PCR^6^, sample testing in the case cohort of <10^6^ copies per sample may not represent a live virus isolate. Unless equipped with a live virus isolate, we cannot be certain whether these retest positive patients were capable of infecting others (contagious) given that they were quarantined at a designated center.

Ninety-three percent of the retest positive patients had mild or moderate severity disease during their first hospitalization. No obvious trending was found in this case series between the initial viral load and first admission symptoms, e.g. temperature (Supplementary Figure 7). This is in line with a previous report that described lack of correlation between transmissibility of COVID-19 and exceeding-positive-threshold-levels of SARS-CoV-2 shedding in the upper respiratory tract^17^. The two readmitted febrile patients with typical clinical manifestations satisfying the first admission criteria may have been capable of transmission given the presence of positivity with both viral load testing and COVID-19 admission symptoms. The key rationale of the local health policy for implementing the described strict center-based quarantine is to prevent transmission. Our findings demonstrate that the effectiveness of this quarantine strategy in the management of the pandemic may be crucial in minimizing late transmission. They also provide insight into the case management of retested positive patients with or without typical COVID-19 symptoms^3,17–31^ and immunity.

The model identified serum concentrations of cholinesterase, calcium, and eGFR as predictors, and elevation of the three markers associated with the risk of recurrence of PCR positivity. Previous studies found that increased levels of cholinesterase, calcium, and eGFR were associated with mild COVID-19^32–34^. In our study, mild or moderate patients more likely to recur with PCR positivity post discharge. Thus, the associations of the blood markers and case outcomes revealed by our model appear to be consistent with previous findings.

Determining those at highest and lowest risks of recurrence of PCR positivity may be an essential component of any strategy to enable readmissions, guide interventions, prevent transmissions, and optimize limited care resource utilization. The analysis described herein builds on our previously validated machine-learning models to predict hospital readmission^35,36^ to develop COVID-19 readmission risk prediction tool^37^. The algorithm described here enables different case management strategies for various risk score thresholds, and facilitates the incorporation of different assumptions about the impact of the intervention and quarantine (Supplementary Figure 1). At discharge, those patients flagged by the algorithm to be at highest risk may need different follow-up or different screening strategies. Most critically, the time and treatment-dependent trajectory to completely clear dormant and/or live SARS-CoV-2 needs to be established.

The driving factors for this model can be collected in any inpatient settings, and therefore it is portable internationally for immediate validation to provide clinical utility in the current COVID-19 pandemic.

## Limitations

This study has several limitations. First, the study population only included COVID-19 patients within a single center in Southern China. Second, the IgM/IgG tests were not performed on all patients due to the late introduction of the antibody test. Thirdly, for model validation, multi-center prospective analyses on additional patients will be required. Fourthly, and most importantly, the (re)infectious ability of SARS-CoV-2 measured positive by the nasopharyngeal swab test needs to be quantified. Live virus isolation from retest positive patients still requires further effort in order to quantify risk associated with recurrence of PCR positivity at a viral RNA level.

## Conclusions

This case series demonstrates that recurrence of SARS-CoV-2 RNA positivity following hospital discharge is relatively common, with a 16.7% rate. Younger patients with less severe index illness were more likely to retest positive. More information will be required to understand the relevance of these findings, which will be critical for informing the management of the pandemic. Our prediction algorithm to identify patients at high risk of recurrence of PCR positivity may facilitate health policy.

## Data Availability

Data are available upon request.

